# Comparison of Influenza Vaccine Effectiveness Estimates Using Test-Negative Prospective Enrollment and Electronic Health Record Data, United States, 2024–2025

**DOI:** 10.64898/2026.07.23.26358258

**Authors:** Ashley M. Price, Callie McLean, Seana Cleary, Aleda M. Leis, Ivana A. Vaughn, Stacey L. House, Sam Ellsworth, Krissy Moehling Geffel, Louise H. Taylor, Manjusha Gaglani, Kempapura Murthy, Elie A. Saade, Christopher Ladikos, Vel Murugan, Joanna L. Kramer, Brian D. Williamson, Erika Kiniry, Emmanuel B. Walter, Natalie AB Bontrager, Sascha R. Ellington, Brendan Flannery, Jessie R. Chung, US Flu VE Network Investigators

**Affiliations:** Influenza Division, US Centers for Disease Control and Prevention, Atlanta, GA, USA; STI Federal, Sault Tribe Incorporated, Sault Ste Marie, Michigan, USA; University of Michigan School of Public Health, Ann Arbor, MI, USA; Henry Ford Health, Detroit, MI, USA; Washington University School of Medicine in St. Louis, Department of Emergency Medicine, St. Louis, MO, USA; University of Pittsburgh School of Medicine, Department of Family Medicine, Pittsburgh, PA, USA; Baylor Scott C White Health, Temple, TX, USA; Baylor College of Medicine, Temple, TX, USA; University Hospitals of Cleveland, Cleveland, OH, USA; Biodesign Center for Personalized Diagnostics and Health Observatory Arizona State University, Tempe, AZ, USA; Phoenix Children’s Hospital, Phoenix, AZ; Kaiser Permanente Washington Health Research Institute, Seattle, WA, USA; Duke Human Vaccine Institute, Duke University School of Medicine, Durham, NC, USA

**Keywords:** Influenza, vaccine effectiveness, test-negative design, electronic medical record

## Abstract

**Background:** Influenza vaccine effectiveness (VE) is assessed annually through prospective enrollment of patients presenting with acute respiratory symptoms in a test-negative study design. Influenza VE has also been estimated from electronic health record (EHR) databases by linking medical diagnoses, laboratory test results, and patient influenza vaccination. There are limited data on agreement between influenza VE estimates from prospective enrollment versus EHR databases.

**Methods:** The US Influenza Vaccine Effectiveness Network prospectively enrolled outpatients meeting clinical screening criteria and collected respiratory specimens to determine influenza virus infection. Seven study sites also identified EHR databases that included diagnostic codes for outpatient encounters associated with medically attended acute respiratory illness (MAARI), clinical respiratory virus testing, and influenza vaccination status. Effectiveness of influenza vaccination against laboratory-confirmed influenza was estimated from both data sources using logistic regression models including patient age, study site, and month of illness as 100(%) x (1 – adjusted odds ratio), comparing influenza vaccination among laboratory-confirmed influenza-positive patients versus laboratory-confirmed influenza-negative patients.

**Results:** From October 2024–April 2025, 2,016 (30%) of 6,793 prospectively enrolled patients and 75,885 (24%) of 282,444 EHR MAARI encounters had laboratory-confirmed influenza virus infection. Effectiveness of vaccination against laboratory-confirmed influenza was 36% (95% confidence interval [CI]: 26-44) among prospectively enrolled patients and 38% (95% CI: 36-39) among EHR MAARI encounters. Comparing influenza VE estimates from the two data sources, confidence intervals overlapped for all age groups except for adults aged ≥65 years: −3% (95% CI: −53–30) among prospective enrollment versus 35% (95% CI: 32–39) VE from EHR MAARI encounters.

**Conclusion:** Overall, influenza VE estimates from retrospective EHR data were similar to VE estimates using the test-negative design with prospective enrollment. The age group-specific differences in estimated VE observed in US adults aged ≥65 years compared with younger age groups merit further investigation.

**Key Points:** Retrospective EHR-based data and prospective enrollment of US patients using a test-negative design produced similar overall estimates of influenza VE during the 2024–2025 season.

## INTRODUCTION

The 2024–2025 influenza season in the United States was the highest severity season since 2017–2018, with an estimated 43 million – 76 million influenza-related illnesses, 19 million – 34 million medical visits, 560,000 – 1.2 million hospitalizations, and 38,000 – 110,000 deaths (1). The US Centers for Disease Control and Prevention (CDC) supports multiple networks to provide annual estimates of influenza vaccine effectiveness (VE) for the prevention of laboratory-confirmed influenza (2). During each influenza season since 2004–2005 (3), the US Influenza Vaccine Effectiveness (Flu VE) Network has prospectively enrolled patients meeting clinical criteria for systematic influenza virus testing. Influenza VE is estimated using a test-negative design (TND) by comparing odds of influenza vaccination among patients who test positive (cases) to those that test negative (controls) for influenza virus infection (4, 5). Since 2021, influenza VE has also been estimated retrospectively using electronic health record (EHR) databases linking medical diagnoses, clinical laboratory test result, and patient influenza vaccination information (6–8). EHR databases include information on large numbers of patients with medically attended acute respiratory infection (MAARI) encounters, providing the ability to rapidly and precisely assess VE against confirmed influenza A and B and product-specific VE. During 2024–2025, influenza VE estimates from EHR databases were consistent with and more precise than estimates from other prospective enrollment test-negative studies (2,9). However, few studies have compared VE estimates from prospective enrollment studies and EHR databases within the same source population. We compared estimates of 2024–2025 influenza VE based on prospective enrollment with retrospective analysis of healthcare databases from US Flu VE Network study sites representing seven geographic areas.

## METHODS

### Study design and population

The study was conducted with participating institutions in the US Influenza Vaccine Effectiveness (Flu VE) Network during the 2024–2025 influenza season: Phoenix Children’s Hospital, Valleywise Health and Arizona State University, Tempe, Arizona; University of Pittsburgh Medical Center (UPMC), Pittsburgh, Pennsylvania; Washington University in St. Louis Affiliated Hospitals and Centers, St. Louis, Missouri; University Hospitals of Cleveland, Ohio; Baylor Scott and White Health, Temple, Texas; University of Michigan Health, Ann Arbor and Henry Ford Health, Detroit, Michigan; and Kaiser Permanente Washington, Seattle, Washington (9). The study protocols were reviewed by CDC, determined to be public health surveillance, and conducted consistent with applicable federal law and CDC policy (45 CFR 46.102(l)(2)). Participants or their parent/guardian provided written or oral consent.

### Prospective enrollment

From October 2024 to April 2025, Flu VE Network staff at outpatient healthcare facilities (comprising primary care, urgent care, and emergency departments) associated with healthcare institutions in Flu VE Network prospective enrollment sites screened and enrolled eligible patients aged ≥8 months who reported acute onset or worsening (≤7 days earlier) of cough. Information collected from enrolled patients included patient age, sex, race/ethnicity, and 2024–2025 seasonal influenza vaccination. At enrollment, study staff collected nasal and/or oropharyngeal swab specimens (only nasal swab specimens were collected for children aged <2 years). Specimens were tested for influenza virus and SARS-CoV-2 infection using real-time reverse-transcription polymerase chain reaction (RT-PCR). In addition, study staff collected documented 2024–2025 influenza vaccination from electronic immunization records.

### Electronic health record (EHR) data

At each study site, study staff extracted electronic health system data on outpatient encounters between October 2024 and May 2025 associated with a pre-defined list of International Classification of Diseases, version 10 (ICD-10) diagnostic codes for outpatient MAARI (**Supplemental Table 1**). Patient medical record numbers and other unique identifiers were used to link MAARI encounters, clinical laboratory test results, and electronic immunization records. Sources of EHR, vaccination and laboratory results for individual patients with MAARI who received healthcare within a defined source population differed by study site. EHR data included outpatient encounters among Kaiser Permanente members in all primary care catchment areas in Washington State. In Arizona, Michigan, Missouri, Ohio, Pennsylvania, and Texas EHR data included outpatient encounters at facilities within provider networks. Multiple MAARI encounters and laboratory results within a +/− 7-day period were considered as a single event. Laboratory-confirmed influenza was defined as one or more positive influenza diagnostic tests within 14 days of the date of initial MAARI encounter, regardless of diagnostic test type. Patients who received a documented dose of any 2024–2025 seasonal influenza vaccine at least two weeks prior to their MAARI encounter were considered vaccinated. To protect patient information, study sites de-identified records before sharing with CDC.

### Data analysis

For each study site, demographic characteristics and influenza test positivity among patients prospectively enrolled in the Flu VE Network using a test-negative design were compared with characteristics and influenza test results among patients with MAARI encounters from EHR data by patient age group, sex and race/ethnicity using Wald chi-squared for categorical variables and nonparametric Wilcoxon rank-sum for continuous variables. We estimated overall and age-specific influenza vaccination coverage with at least one documented dose of any 2024–2025 seasonal influenza vaccine and compared age-specific percentages among influenza test-negative patients enrolled in the Flu VE Network to EHR MAARI encounters with negative influenza diagnostic tests using chi-squared tests (**Supplemental Table 2**). Patients with a positive SARS-CoV-2 test were excluded from both analyses. Information on home-based testing was collected only among patients prospectively enrolled in the Flu VE Network and was not available for EHR MAARI encounters. Adjusted odds ratios and 95% confidence intervals (CI) were estimated using logistic regression models controlling for participant age, study site, and month of illness onset. To determine agreement between estimates, we assessed whether point estimates from the EHR analysis fell within the 95% confidence limits of the Flu VE Network prospective enrollment analysis during the 2024–2025 influenza season. VE against influenza was estimated as 100 (%) x (1-adjusted Odds Ratios [aOR]) comparing vaccination status by influenza test result. For the Flu VE Network analysis, cases were defined as prospectively enrolled patients with a positive influenza test, and controls were defined as prospectively enrolled patients with a negative influenza test. For the EHR analysis, cases were defined as patients with a MAARI encounter and a positive clinical influenza test, and controls were defined as patients with a MAARI encounter and a negative clinical influenza test. We conducted analyses using R (v4.4.0; R Core Team 2024) (10).

## RESULTS

Overall, 6,793 patients meeting clinical screening criteria were prospectively enrolled in the Flu VE Network; 2,016 (30%) had RT-PCR confirmed influenza virus infection (**Table 1**). Influenza positivity among prospectively enrolled Flu VE Network patients ranged from 11% to 39% by site (**Supplemental Figure 1**). During the same period, there were 282,444 MAARI encounter patients with associated influenza diagnostic test results from EHR data; 67,532 (24%) had ≥1 positive influenza clinical tests for the encounter. Forty percent of MAARI encounters were enrolled at US Flu VE prospective enrollment clinics. The number of MAARI encounters included varied by study site, ranging from 16,464 in Pennsylvania to 77,079 in Missouri (**Table 1**). Influenza positivity among those tested for influenza in the EHR database ranged from 16% to 30% by site (**Supplemental Figure 1**).

**Table 1.**
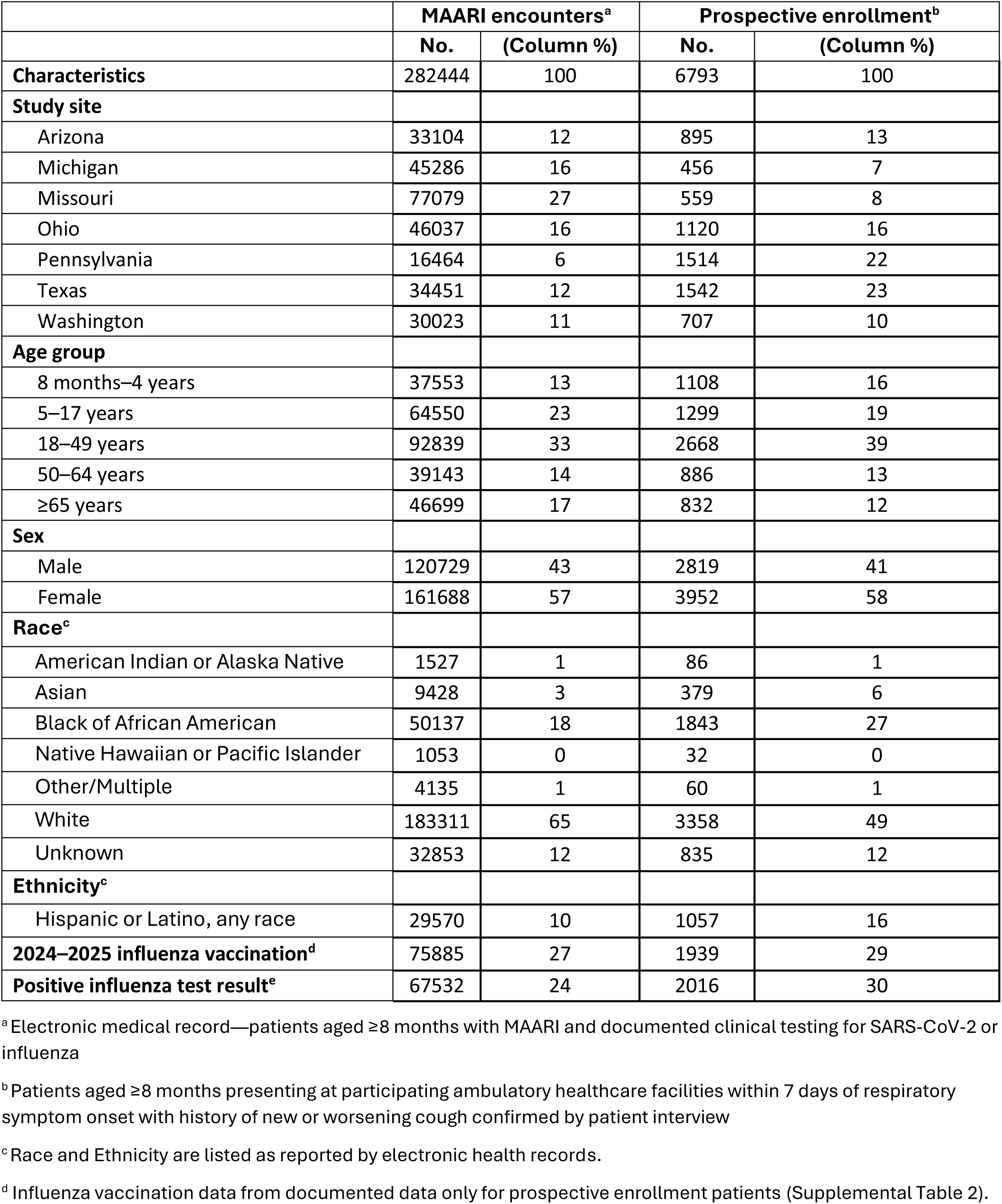
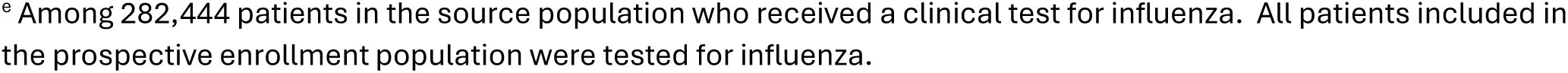
Comparison of patients with electronic health record of medically attended acute respiratory illness-associated encounter versus patients prospectively enrolled in the US Influenza Vaccine Effectiveness Network during the 2024–2025 influenza season.

|  | **MAARI encounters<sup>a</sup>** |  | **Prospective enrollment<sup>b</sup>** |  |
| --- | --- | --- | --- | --- |
|  | **No.** | **(Column %)** | **No.** | **(Column %)** |
| **Characteristics** | 282444 | 100 | 6793 | 100 |
| **Study site** |  |  |  |  |
| Arizona | 33104 | 12 | 895 | 13 |
| Michigan | 45286 | 16 | 456 | 7 |
| Missouri | 77079 | 27 | 559 | 8 |
| Ohio | 46037 | 16 | 1120 | 16 |
| Pennsylvania | 16464 | 6 | 1514 | 22 |
| Texas | 34451 | 12 | 1542 | 23 |
| Washington | 30023 | 11 | 707 | 10 |
| **Age group** |  |  |  |  |
| 8 months–4 years | 37553 | 13 | 1108 | 16 |
| 5–17 years | 64550 | 23 | 1299 | 19 |
| 18–49 years | 92839 | 33 | 2668 | 39 |
| 50–64 years | 39143 | 14 | 886 | 13 |
| ≥65 years | 46699 | 17 | 832 | 12 |
| **Sex** |  |  |  |  |
| Male | 120729 | 43 | 2819 | 41 |
| Female | 161688 | 57 | 3952 | 58 |
| **Race<sup>c</sup>** |  |  |  |  |
| American Indian or Alaska Native | 1527 | 1 | 86 | 1 |
| Asian | 9428 | 3 | 379 | 6 |
| Black or African American | 50137 | 18 | 1843 | 27 |
| Native Hawaiian or Pacific Islander | 1053 | 0 | 32 | 0 |
| Other/Multiple | 4135 | 1 | 60 | 1 |
| White | 183311 | 65 | 3358 | 49 |
| Unknown | 32853 | 12 | 835 | 12 |
| **Ethnicity<sup>c</sup>** |  |  |  |  |
| Hispanic or Latino, any race | 29570 | 10 | 1057 | 16 |
| **2024–2025 influenza vaccination<sup>d</sup>** | 75885 | 27 | 1939 | 29 |
| **Positive influenza test result<sup>e</sup>** | 67532 | 24 | 2016 | 30 |

<sup>a</sup>Electronic medical record—patients aged ≥8 months with MAARI and documented clinical testing for SARS-CoV-2 or influenza
<sup>b</sup>Patients aged ≥8 months presenting at participating ambulatory healthcare facilities within 7 days of respiratory symptom onset with history of new or worsening cough confirmed by patient interview
<sup>c</sup>Race and Ethnicity are listed as reported by electronic health records.
<sup>d</sup>Influenza vaccination data from documented data only for prospective enrollment patients (Supplemental Table 2).
<sup>e</sup> Among 282,444 patients in the source population who received a clinical test for influenza. All patients included in the prospective enrollment population were tested for influenza.

Sex, age group, and racial/ethnic demographics were similar among enrolled patients and patients included in EHR MAARI encounters. At most sites, MAARI patients were more likely to be female, making up 48%–63% of encounters (**Supplemental Figure 2**). Age group distributions were similar across most sites. Pediatric patients aged 8 months to 8 years accounted for 36% of all MAARI patients with 36% identified in Arizona. In Pennsylvania, 47% of enrolled patients were <18, while in Missouri, 78% were ≥18 (**Supplemental Figure 3**). Across all sites, most patients included in both the EHR MAARI encounters and prospectively enrolled population identified as White except in Missouri where most prospectively enrolled identified as Black **(Supplemental Figure 4**). Black patients accounted for ≤20% of MAARI encounters except in Missouri (25%), while Asian patients accounted for 12% in Washington state. Percentages of patients who identified as Hispanic or Latino were higher among prospectively enrolled patients compared to EHR MAARI patient encounters (**Table 1**).

Among 6,793 prospectively enrolled patients, 1,939 (29%) had documented 2024–2025 influenza vaccination, compared with 75,885 (27%) of patients with EHR MAARI encounters (**Table 2**). Percentages with documented vaccination were highest in Washington state (47% and 40% among prospectively enrolled patients and MAARI encounters, respectively) and lowest in Arizona (11% and 8%). Patterns in age group-specific influenza vaccination were similar among patients with MAARI encounters and prospectively enrolled patients at each study site: among children aged 8 months through 17 years, average coverage was 23% among prospectively enrolled patients versus 21% among children with MAARI encounters (**Supplemental Figure 5**). Average influenza vaccination coverage among adults aged ≥18 years was 33% among prospectively enrolled patients and 36% among adults with MAARI encounter. In addition, age group-specific percentages of influenza vaccination agreed with national immunization survey data for the seven states with Flu VE Network study sites (**Supplemental Figure 5**). Among prospectively enrolled patients, 2024–2025 influenza vaccination rates were lower among Black than White patients (20% vs. 34%). Similarly, among patients included in EHR MAARI encounters, vaccination rates were lower among Black than White patients (19% and 30%).

**Table 2.** Comparison of electronic immunization record documentation of 2024–2025 influenza vaccination among patients with MAARI encounter and negative clinical influenza test results versus influenza test-negative patients prospectively enrolled in the US Influenza Vaccine Effectiveness Network study.

|  | MAARI encounters<sup>a</sup> |  |  | Prospective enrollment<sup>b</sup> |  |  |
| --- | --- | --- | --- | --- | --- | --- |
|  | Vaccinated | Total | Row % | Vaccinated | Total | Row % |
| **Overall** | 75885 | 282444 | 27 | 1939 | 6793 | 29 |
| **Study sites** |  |  |  |  |  |  |
| Arizona | 2555 | 33104 | 8 | 97 | 895 | 11 |
| Michigan | 12958 | 45286 | 29 | 211 | 456 | 46 |
| Missouri | 24645 | 77079 | 32 | 92 | 559 | 16 |
| Ohio | 11899 | 46037 | 26 | 135 | 1120 | 12 |
| Pennsylvania | 4184 | 16464 | 25 | 536 | 1514 | 35 |
| Texas | 7688 | 34451 | 22 | 536 | 1542 | 35 |
| Washington | 11956 | 30023 | 40 | 332 | 707 | 47 |
| **Age group** |  |  |  |  |  |  |
| 8 months–4 years | 8048 | 37553 | 23 | 327 | 1108 | 30 |
| 5–17 years | 11057 | 64550 | 18 | 273 | 1299 | 21 |
| 18–49 years | 17390 | 92839 | 19 | 532 | 2668 | 20 |
| 50–64 years | 12921 | 39143 | 34 | 327 | 886 | 37 |
| ≥65 years | 24652 | 46699 | 54 | 480 | 832 | 58 |
| **Sex** |  |  |  |  |  |  |
| Male | 29945 | 120729 | 25 | 751 | 2819 | 27 |
| Female | 45934 | 161688 | 29 | 1181 | 3952 | 30 |
| **Race<sup>c</sup>** |  |  |  |  |  |  |
| American Indian or Alaska Native | 377 | 1527 | 25 | 22 | 86 | 26 |
| Asian | 3575 | 9428 | 38 | 128 | 379 | 34 |
| Black or African American | 9657 | 50137 | 19 | 376 | 1843 | 20 |
| Other/Multiple | 295 | 1053 | 28 | 8 | 32 | 25 |
| Native Hawaiian or Pacific Islander | 1249 | 4135 | 30 | 12 | 60 | 20 |
| White | 55588 | 183311 | 30 | 1120 | 3358 | 34 |
| Unknown | 5144 | 32853 | 16 | 173 | 835 | 21 |
| **Ethnicity<sup>c</sup>** |  |  |  |  |  |  |
| Hispanic or Latino, any race | 4992 | 29570 | 17 | 239 | 1057 | 23 |

<sup>a</sup> Source population included electronic health records for patients aged ≥8 months with MAARI and documented clinical testing for influenza with a negative result.
<sup>b</sup> Patients aged ≥8 months presenting at participating ambulatory healthcare facilities within 7 days of respiratory symptom onset with history of new or worsening cough confirmed by patient interview
<sup>c</sup> Persons identifying as Hispanic might have been of any race.

After excluding 20,901 EHR MAARI encounters with positive SARS-CoV-2 test results, 214,912 EHR MAARI encounters with negative influenza test results were included in the comparison population for influenza vaccine effectiveness. Overall estimates of 2024–2025 vaccine effectiveness against laboratory confirmed influenza were similar in the two data sources (**Figure 1**). Among all patients, VE estimated from patient MAARI encounters was 38% (CI: 36 to 39) versus 36% (CI: 26 to 44) among prospectively enrolled patients. Among children aged 8 months to 4 years, VE estimated from patient MAARI encounters was 47% (95% CI: 43–51) versus 51% (95% CI: 29–66) among prospectively enrolled patients. However, among adults aged ≥65 years, VE was 35% (95% CI: 31– 39) based on patient MAARI encounters but was not statistically significant (−3%; 95% CI: −53 to 30) among prospectively enrolled patients.

**Figure 1.**
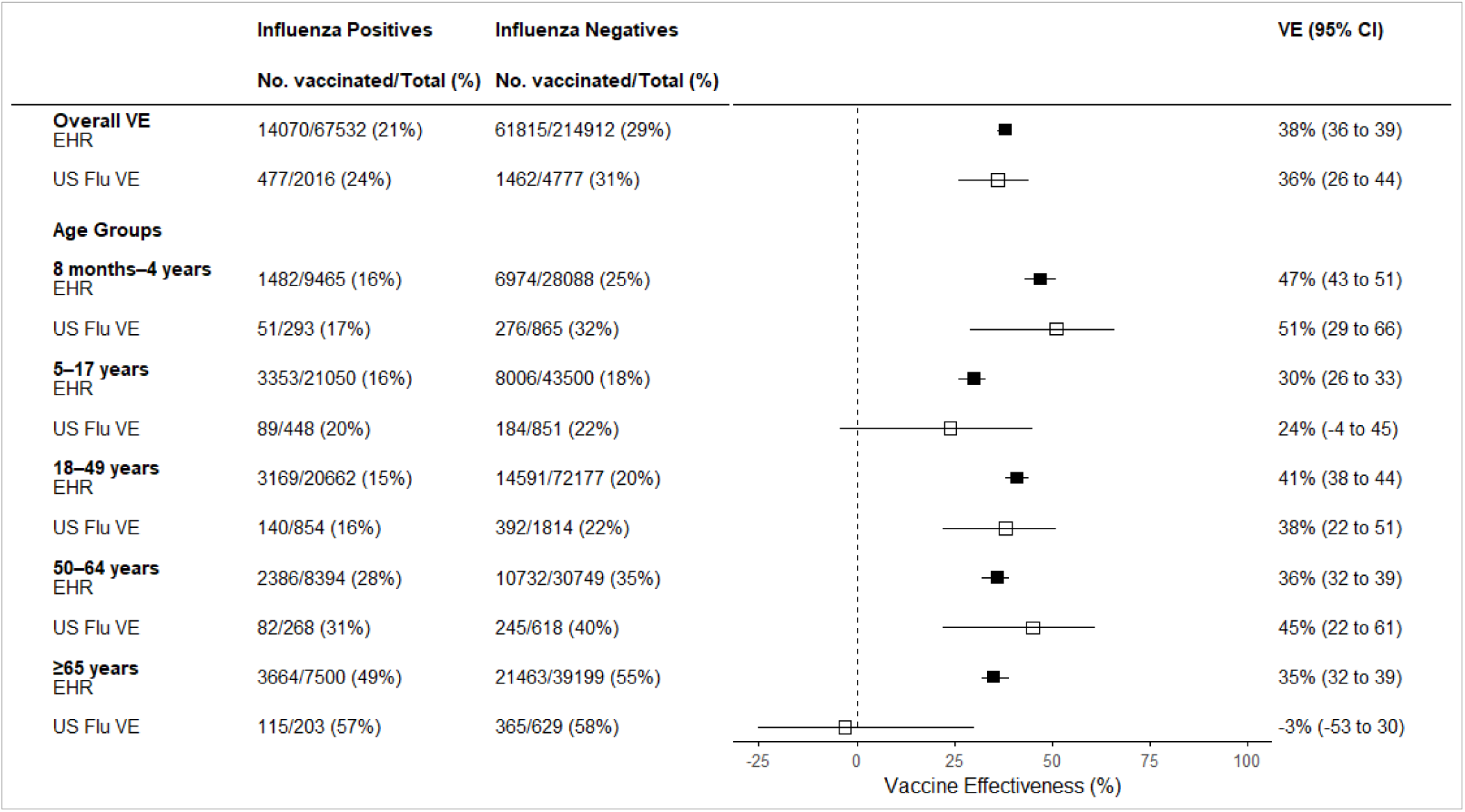
**Influenza** vaccine effectiveness (VE) against laboratory confirmed influenza based on medically attended acute respiratory illness encounters using electronic health record (EHR) databases versus patients prospectively enrolled in the Influenza VE Network, 2024–2025. Vaccine effectiveness was estimated with logistic regression adjusting for age group, race/ethnicity, month of illness, and study site. Boxes show point estimates with 95% confidence intervals. Concordance was defined as US Flu VE point estimates within 95% confidence bounds for EHR database estimates.

## DISCUSSION

Findings suggest that estimates of influenza vaccine effectiveness against confirmed influenza during the US 2024–2025 influenza season based on retrospective EHR data were generally consistent with influenza VE estimates using the commonly utilized systematically screened and prospectively enrolled test-negative design (TND). Both approaches produced similar, statistically significant VE point estimates for protection against laboratory-confirmed influenza-associated outpatient visits among children and adults aged <65 years. However, among patients aged ≥65 years enrolled in the US Flu VE network, influenza VE estimates were not statistically significant. The EHR estimates from our study were consistent with other published EHR-based estimates for this age group (31%–44%) (11–12).

EHR database analyses from these seven participating sites and affiliated healthcare systems were similar to previously published estimates of protection against laboratory-confirmed influenza-associated emergency department and urgent care visits from multiple large healthcare networks for 2024–2025 Influenza season in the United States (12). With increased use of SARS-CoV-2 and influenza diagnostic tests being ordered for patients presenting with uncomplicated respiratory illness in urgent care clinics and emergency departments, EHR data may provide opportunities to estimate annual influenza VE against laboratory-confirmed influenza (13). Linkage of clinical test results, including both positive and negative results, with diagnostic codes and vaccination records allows retrospective analyses of VE against laboratory-confirmed outcomes using patient MAARI encounters associated influenza diagnostic testing. However, with changes possible in clinical diagnostic testing practices and variable sensitivity and specificity of clinical influenza tests, TND studies with systematic screening and prospective enrollment of patients meeting clinical criteria and testing for influenza virus infection with highly sensitive and specific RT-PCR continue to be important. Studies that screen and enroll patients meeting clinical criteria like the US Flu VE Network include patient contact to gain information not routinely found in electronic health records about reported individual symptoms, date of illness onset, perceived severity of illness, individual factors such as smoking habits, and behaviors such as at-home testing practices (13). They also provide an opportunity for follow-up interactions to collect additional information like illness duration, productivity loss due to illness, and secondary transmission within households. Further, active enrollment studies can systematically collect specimens such as sera and peripheral blood mononuclear cells that provide additional insights into protection afforded by vaccines (14). These traditional TND studies also allow for the ability to produce estimates of VE by influenza A subtype and genetic groups which can inform annual influenza vaccine strain selection (15). Comparison of prospective enrollment and patient MAARI encounters in EHR data also found similar proportions of patients with documented receipt of 2024–2025 influenza vaccination. US Flu VE Network provides information on patient characteristics associated with influenza vaccination, as well as patient reported vaccination to assess completeness of electronic immunization records. US Flu VE network sites also use vaccination verification methods reconciling external immunizations using state immunization registries and medical release of information forms. While differences between vaccination coverage by study site may be associated with completeness of electronic records, geographic trends by age group were consistent with national immunization surveys. Influenza vaccination coverage remains well below the Healthy People 2030 target of 70% vaccination (16).

An important limitation of any comparison of VE estimates from observational studies is the lack of a reference estimate. To be conservative when assessing agreement between estimates, we assessed whether point estimates from the EHR analysis fell within the confidence limits of the Flu VE Network prospective enrollment analysis during the 2024–2025 influenza season. Secondly, although retrospective analyses of EHR medical encounters with associated influenza testing and traditional TND studies reduce potential bias associated with differences in healthcare seeking behavior (17, 18), biases due to other health behaviors including use of at-home rapid diagnostic tests may have affected VE estimates using both approaches (19, 20). Thirdly, source populations for MAARI encounters were defined differently by study sites. In addition, some enrolled patients may have been included in the MAARI source population if they received clinical SARS-CoV-2 or influenza tests. While both analyses included patients with ARI who sought healthcare, prospectively enrolled patients in the US Flu VE Network consented to swab collection and completed enrollment questionnaires. Therefore, enrolled patients were less representative of the source population than all patients tested for SARS-CoV-2 or influenza.

In conclusion, retrospective analyses of large EHR databases and test-negative design with prospective enrollment of US patients meeting symptom criteria produced overall concordant estimates of influenza vaccine effectiveness during one high-severity influenza season. Findings support use of large EHR databases with linked clinical test results to measure seasonal influenza VE in settings with widespread clinical testing and documentation of influenza vaccination. Changes in clinical testing practices, health insurance reimbursement and guidance for testing to inform clinical management (21) may influence frequency of testing and completeness of electronic databases with linked clinical test results. The age group-specific differences in estimated VE observed in adults aged ≥65 years compared with younger age groups merit further investigation. Comparisons of the two approaches in the same source population over multiple seasons with different circulating viruses and levels of vaccine effectiveness may inform use of EHR databases for continued estimation of seasonal VE against influenza and other respiratory diseases.

## Data Availability

All data produced in the present study are available upon reasonable request to the authors.

## Disclaimer

The findings and conclusions in this report are those of the authors and do not necessarily represent the official position of the Centers for Disease Control and Prevention.

## US Flu VE Investigators

Emily Martin (University of Michigan); Benjamin Rapp, Jeremy Russell, and Joshua Jackson (Washington University School of Medicine in St. Louis); Gabriella M. Alicea, Donna J. Stoliker, Richard K. Zimmerman, Mary Patricia Nowalk, and Eugene M. Sadhu (University of Pittsburgh); Jason Ramm, Tresa McNeal, Manohar Mutnal, Chandni Raiyani, Mufaddal Mamawala, Spencer Rose, and Michael Smith (Baylor Scott C White Health); Zainab Albar, Alexia El Khoury, and Joy Abou Farah (University Hospitals of Cleveland); Kelly Conard and Shodhan Manda (Biodesign Center for Personalized Diagnostics, Arizona State University, Tempe, AZ); Sean Reed and Mehal Patel (Phoenix Children’s Hospital, Phoenix, AZ); Lora Nordstrom and Raquel Salgado (Valleywise Health, Phoenix, AZ); Rachael Doud and C. Hallie Phillips (Kaiser Permanente Washington Health Research Institute); Olivia Williams and Darren Morrow (Duke University).

---

**Supplemental Table 1.** International Classification of Diseases, version 10 (ICD-10) diagnostic codes used to identify patients with medically attended acute respiratory illness (MAARI).

| ICD-10 code | Condition |
| --- | --- |
| J00 | Acute nasopharyngitis [common cold] |
| J01.* | Acute sinusitis |
| J02.* | Acute pharyngitis |
| J03.* | Acute tonsillitis |
| J04.* | Acute laryngitis and tracheitis |
| J05.* | Acute obstructive laryngitis [croup] and epiglottitis |
| J06.* | Acute upper respiratory infections of multiple and unspecified sites |
| J20.* | Acute bronchitis |
| J21.* | Acute bronchiolitis |
| J22.* | Unspecified acute lower respiratory infection |
| B97.4* | Respiratory syncytial virus as the cause of diseases classified elsewhere |
| J40.* | Bronchitis, not specified as acute or chronic |
| J41.* | Simple and mucopurulent chronic bronchitis |
| J42.* | Unspecified chronic bronchitis |
| J44.* | Other chronic obstructive pulmonary disease |
| J45.* | Asthma |
| J43.* | Emphysema |
| H66.0* | Acute suppurative otitis media |
| H66.4* | Suppurative otitis media, unspecified |
| H66.9* | Otitis media, unspecified |
| H67.* | Otitis media in diseases classified elsewhere |
| R06.2 | Wheezing |
| R50.9 | Fever, unspecified |
| R50.81* | Fever presenting with conditions classified elsewhere |
| R53.81* | Other malaise |
| R53.83* | Other fatigue |
| R68.83 | Chills (without fever) |
| A37.01 | Whooping cough due to *Bordetella pertussis* with pneumonia |
| A37.11 | Whooping cough due to Bordetella parapertussis with pneumonia |
| A37.81 | Whooping cough due to other Bordetella species with pneumonia |
| A37.91 | Whooping cough, unspecified species with pneumonia |
| B25.0 | Cytomegaloviral pneumonitis |
| J09.* | Influenza due to certain identified influenza viruses |
| J10.* | Influenza due to other identified influenza virus |
| J11.* | Influenza due to unidentified influenza virus |
| J12.* | Viral pneumonia, not elsewhere classified |
| J13.* | Pneumonia due to Streptococcus pneumoniae |
| J14.* | Pneumonia due to Hemophilus influenzae |
| J15.* | Bacterial pneumonia, not elsewhere classified |
| J16.* | Pneumonia due to other infectious organisms, not elsewhere classified |
| J17.* | Pneumonia in diseases classified elsewhere |
| J18.* | Pneumonia, unspecified organism |
| R05 | Cough |
| R06.00* | Dyspnea unspecified |
| R06.02* | Shortness of breath |
| R06.1* | Stridor |
| R06.82* | Tachypnea, not elsewhere classified |
| J39.8* | Other specified diseases of upper respiratory tract |
| J39.9* | Disease of upper respiratory tract, unspecified |
| B34.2 | Coronavirus infection, unspecified |
| J12.89 | Pneumonia due to Wuhan coronavirus |
| B97.29* | Other coronavirus as the cause of diseases classified elsewhere |
| R68.89 | Suspected Wuhan coronavirus infection, but also “Other general symptoms and signs” |
| Z20.828 | Exposure to Wuhan coronavirus |
| Z03.818* | COVID-19 Test ordered/pending and clinically unsure |
| U07.1* | Patients clinically suspected to have COVID-19 illness or asymptomatic COVID-19 test-positive patients |

\*Includes all sub-codes

**Supplemental Table 2.**
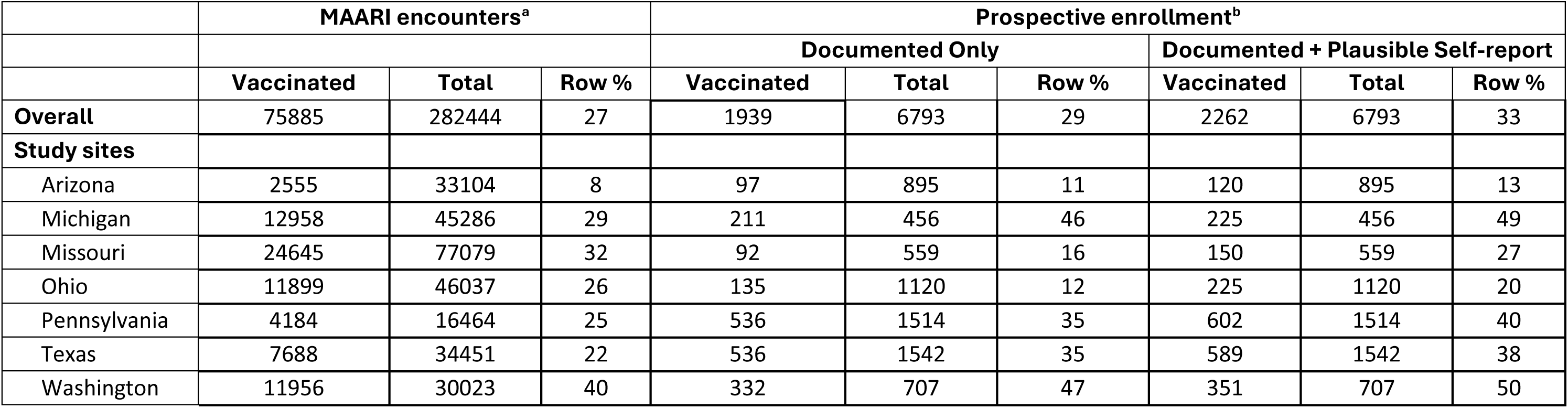
Comparison of documented and self-reported influenza vaccination status among test-negative patients with medically attended acute respiratory illness encounters identified through electronic health records (EHR) versus prospectively enrolled test-negative patients in the US Flu VE Network, 2024–2025 influenza season.

**Supplemental Figure 1.**
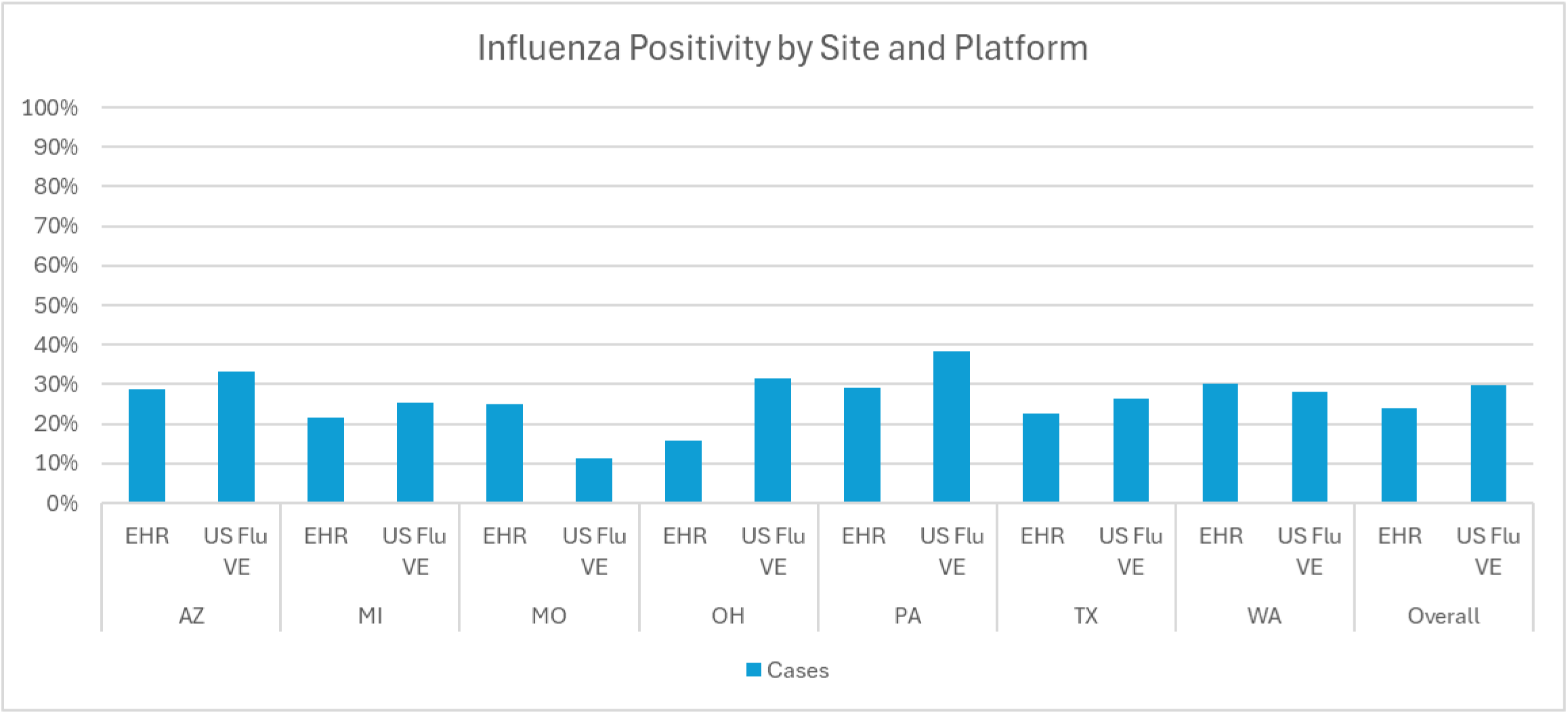
Distributions by influenza test results of patients with medically attended acute respiratory illness in participating healthcare system electronic health records (EHR) versus prospectively enrolled patients in US Flu VE Network, 2024–2025 influenza season. Percentages include all medically attended acute respiratory illness encounters and prospectively enrolled Flu VE patients regardless of respiratory virus test result or influenza vaccination status.

**Supplemental Figure 2.**
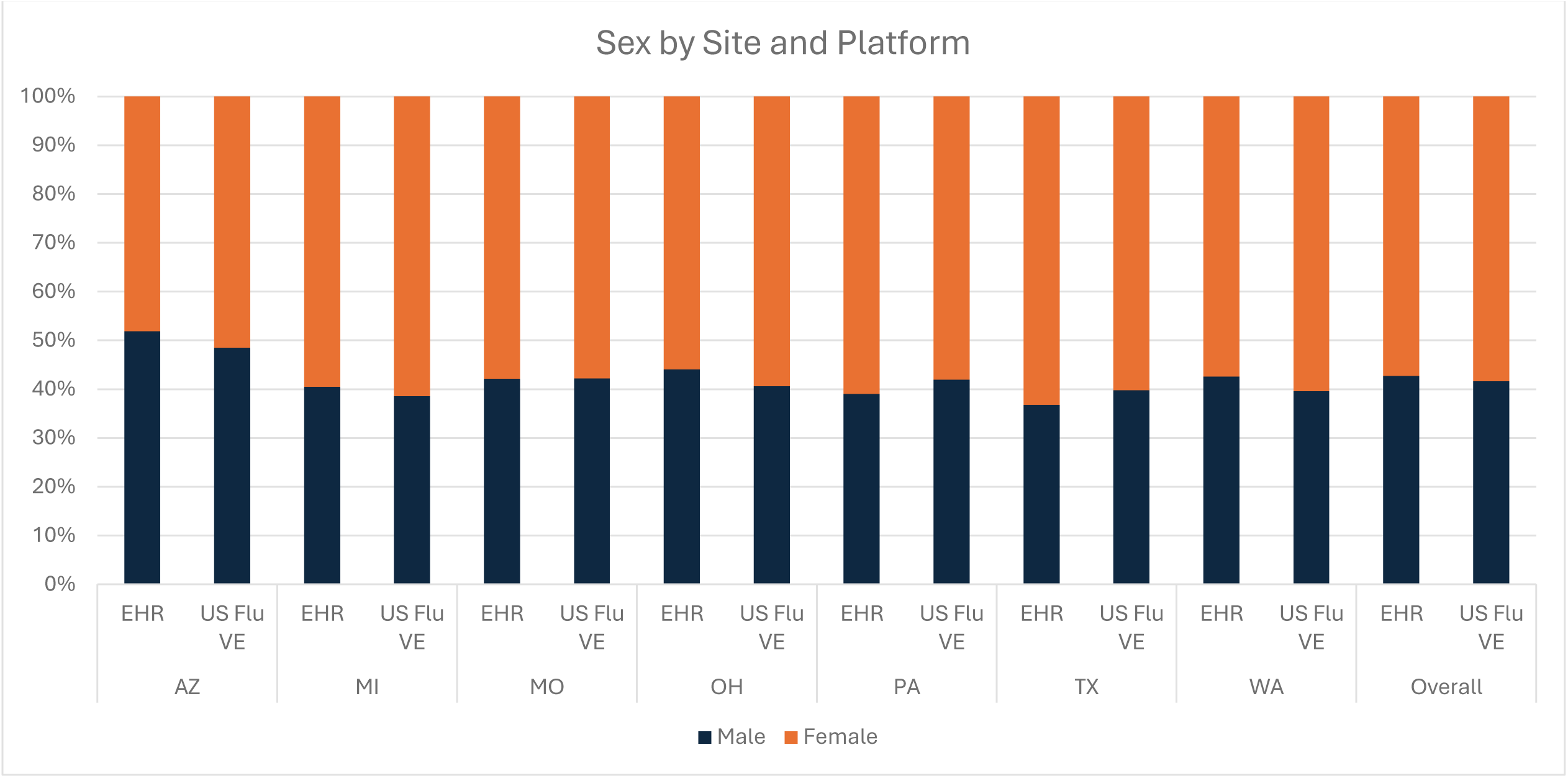
Distributions by sex of patients with medically attended acute respiratory illness in participating healthcare system electronic health records (EHR) versus prospectively enrolled patients in US Flu VE Network, 2024–2025 influenza season. Percentages include all medically attended acute respiratory illness encounters and prospectively enrolled Flu VE Network patients regardless of respiratory virus test result or influenza vaccination status.

**Supplemental Figure 3.**
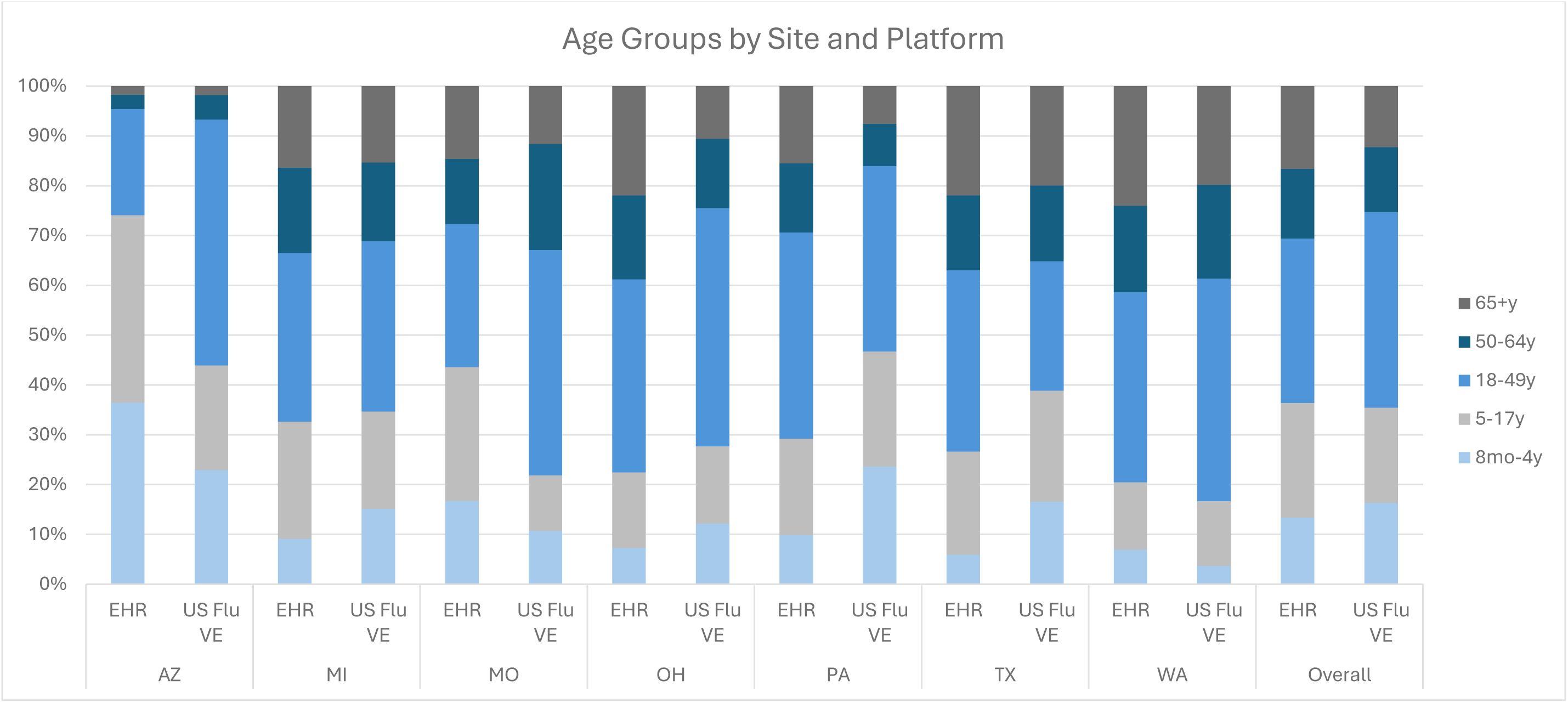
Distributions by age group of patients with medically attended acute respiratory illness in participating healthcare system electronic health records (EHR) versus prospectively enrolled patients in US Flu VE Network, 2024–2025 influenza season. Percentages include all medically attended acute respiratory illness encounters and prospectively enrolled Flu VE Network patients regardless of respiratory virus test result or influenza vaccination status.

**Supplemental Figure 4.**
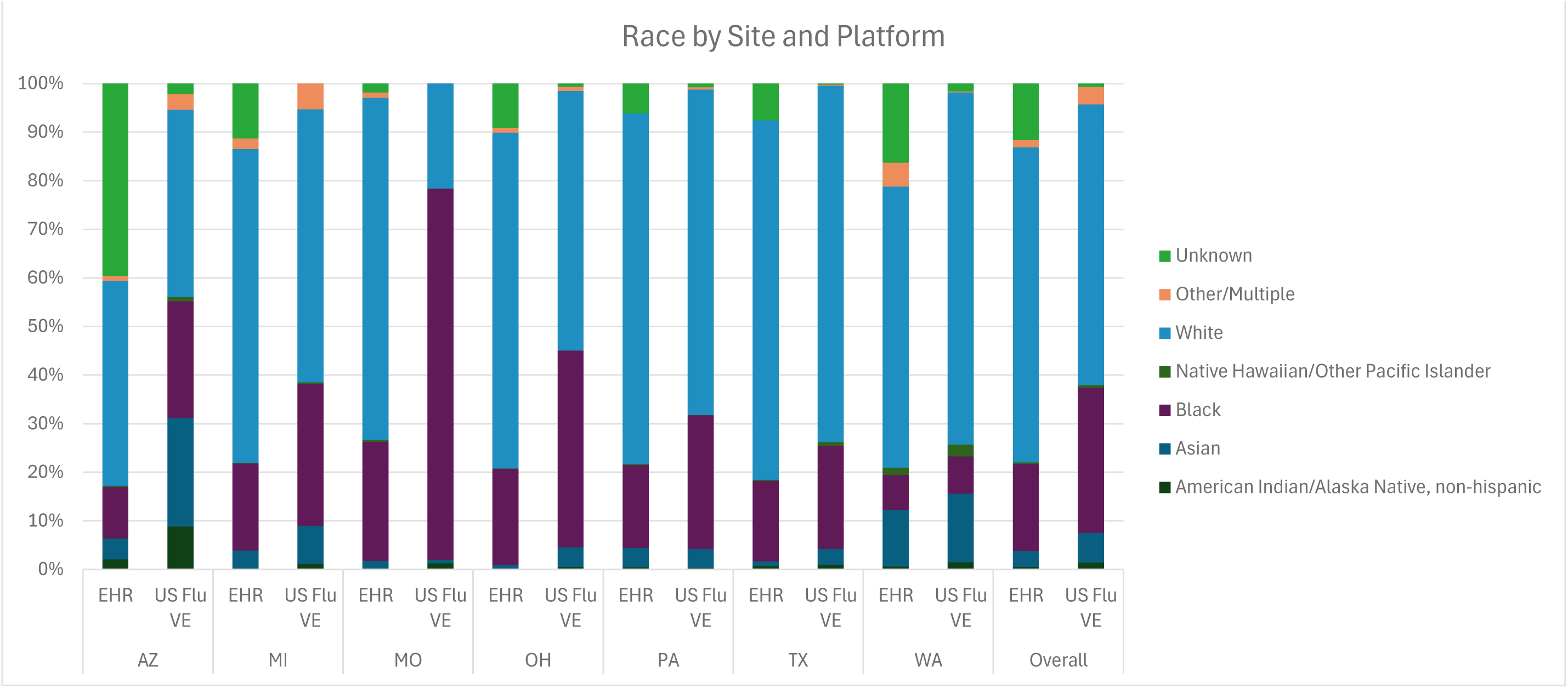
Distributions by race of patients with medically attended acute respiratory illness in participating healthcare system electronic health records (EHR) versus prospectively enrolled patients in the Flu VE Network, 2024–2025. Percentages include all medically attended acute respiratory illness encounters and prospectively enrolled Flu VE Network patients regardless of respiratory virus test result or influenza vaccination status.

**Supplemental Figure 5.**
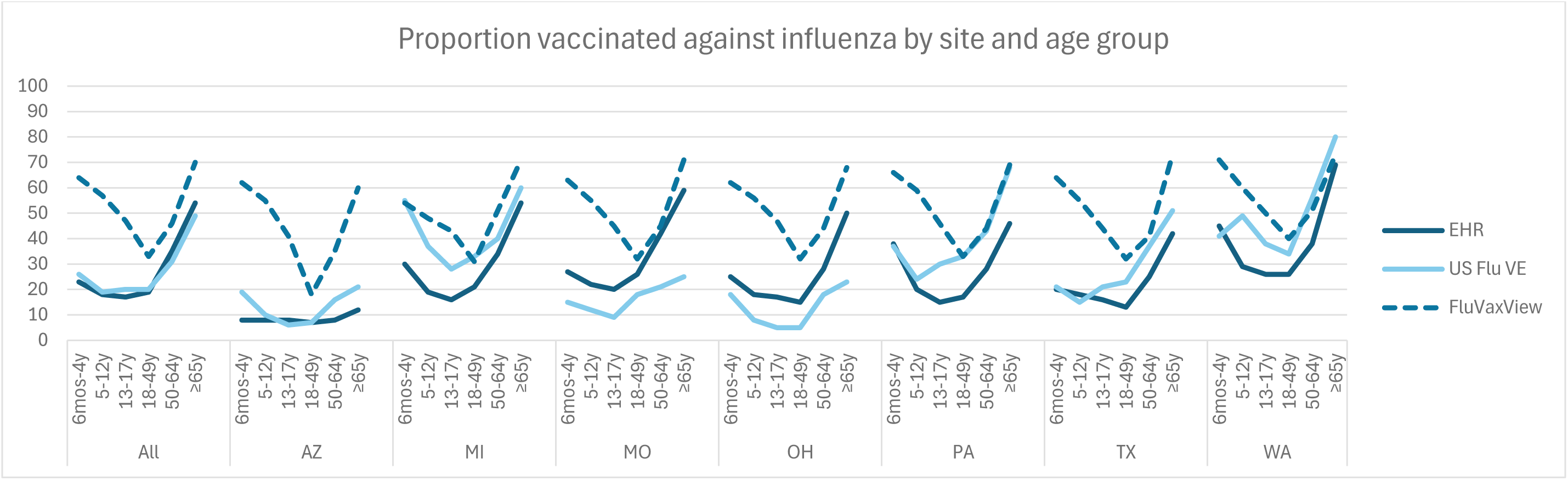
Influenza vaccination percentages by age group and study site among patients with medically attended acute respiratory illness and negative influenza tests in participating healthcare system electronic health records (EHR)^a^ versus prospectively enrolled patients in Flu VE Network^b^, and state influenza vaccination coverage estimates^c^, 2024–2025 influenza season. **^a^** Documented influenza vaccination for medically attended acute respiratory illness encounters associated with negative influenza tests in participating healthcare systems **^b^** Documented influenza vaccination among prospectively enrolled, influenza test-negative patients aged 6 months or older at 7 US Flu VE Network sites **^c^** Centers for Disease Control and Prevention, Influenza Vaccination Coverage for Person 6 months and Older at the state level https://www.cdc.gov/fluvaxview/interactive/index.html

## References

1. Centers for Disease Control and Prevention. 2024-2025 Influenza Season Summary: Severity, Disease Burden, and Burden Prevented. Flu Burden. Centers for Disease Control and Prevention; 2025 [Available from: https://www.cdc.gov/flu-burden/php/data-vis-vac/2024-2025-prevented.html].

2. Frutos AM, Cleary S, Reeves EL, et al. Interim Estimates of 2024-2025 Seasonal Influenza Vaccine Effectiveness - Four Vaccine Effectiveness Networks, United States, October 2024-February 2025. MMWR Morb Mortal Wkly Rep. 2025;74:83–90.

3. Centers for Disease Control and Prevention. Flu Vaccines Work: Centers for Disease Control and Prevention; [Available from: https://www.cdc.gov/flu-vaccines-work/index.html].

4. Foppa IM, Haber M, Ferdinands JM, Shay DK. The case test-negative design for studies of the effectiveness of influenza vaccine. Vaccine. 2013;31(30):3104–9.

5. Jackson ML, Nelson JC. The test-negative design for estimating influenza vaccine effectiveness. Vaccine. 2013;31(17):2165–8.

6. Tenforde MW, Weber ZA, DeSilva MB, Stenehjem E, Yang DH, Fireman B, et al. Vaccine Effectiveness Against Influenza-Associated Urgent Care, Emergency Department, and Hospital Encounters During the 2021-2022 Season, VISION Network. J Infect Dis. 2023;228(2):185–95.

7. Tenforde MW, Weber ZA, Yang DH, DeSilva MB, Dascomb K, Irving SA, et al. Influenza Vaccine Effectiveness Against Influenza A-Associated Emergency Department, Urgent Care, and Hospitalization Encounters Among US Adults, 2022-2023. J Infect Dis. 2024;230(1):141–51.

8. Tenforde MW, Reeves EL, Weber ZA, Tartof SY, Klein NP, Dascomb K, et al. Influenza vaccine effectiveness against hospitalizations and emergency department or urgent care encounters for children, adolescents, and adults during the 2023-2024 season, United States. Clin Infect Dis. 2025;81(3):667–78.

9. Chung JR, Price AM, House SL, Mills J, Wernli KJ, Sanchez M, et al. Influenza vaccine effectiveness against outpatient acute respiratory illness with laboratory-confirmed influenza, United States, 2024-25 season. medRxiv. 2026 [Available from: 10.64898/2026.03.24.26348229.

10. R Foundation for Statistical Computing. R: A language and environment for statistical computing Vienna, Austria [Available from: https://www.R-project.org/.

11. Zhu S, Quint J, León TM, et al. Influenza Vaccine and Associated Infection and Death in California, 2024 to 2025. JAMA Netw Open. 2026;9(6):e2617684. doi:10.1001/jamanetworkopen.2026.17684

12. DeCuir J, Reeves EL, Weber ZA, Yang DH, Irving SA, Tartof SY, et al. Influenza vaccine effectiveness against influenza-associated hospitalizations and emergency department or urgent care encounters among children and adults — United States, 2024–25 season. medRxiv. 2026 [Available from: 10.64898/2026.04.22.26350853].

13. Toepfer AP, Rutkowski RE, Sahni LC, et al. Clinical Testing for COVID-19, Influenza, and RSV in Hospitalized Youths, 2016-2024. JAMA Netw Open. 2025;8(9):e2531499. Published 2025 Sep 2. doi:10.1001/jamanetworkopen.2025.31499

14. Okoli GN, Cowling BJ. Improved Methods for Vaccine Effectiveness Studies. J Infect Dis. 2025;231(6):1367–70.

15. World Health Organization. Recommendations announced for influenza vaccine composition for the 2024–2025 northern hemisphere influenza season. Geneva: World Health Organization; 2024. Available from: https://www.who.int/news/item/[2024-2025-page]

16. Office of Disease Prevention and Health Promotion. Increase the proportion of people who get a flu vaccine every year — IID-09 [Internet]. U.S. Department of Health and Human Services; [cited 2026 Apr 21]. [Available from: https://odphp.health.gov/healthypeople/objectives-and-data/browse-objectives/vaccination/increase-proportion-people-who-get-flu-vaccine-every-year-iid-09].

17. Sullivan SG, Feng S, Cowling BJ. Potential of the test-negative design for measuring influenza vaccine effectiveness: a systematic review. Expert Rev Vaccines. 2014;13(12):1571–91.

18. Sullivan SG, Tchetgen EJ, Cowling BJ. Theoretical Basis of the Test-Negative Study Design for Assessment of Influenza Vaccine Effectiveness. Am J Epidemiol. 2016;184(5):345–53.

19. Qasmieh SA, Ferdinands JM, Chung JR, Wiegand RE, Flannery B, Rane MS, et al. Magnitude of potential biases in COVID-19 vaccine effectiveness studies due to differential health care seeking following home testing: implications for test-negative design studies. Am J Epidemiol. 2026;kwag021. doi:10.1093/aje/kwag021.

20. van Hagen CCE, Vos ERA, Delaunay CL, de Melker HE, Kissling E, Knol MJ. The effect of SARS-CoV-2 testing on healthcare seeking behaviour at primary care level: implications for COVID-19 vaccine effectiveness estimates in test-negative design studies. medRxiv. 2025:2025.03.31.25324928.

21. Uyeki TM, Bernstein HH, Bradley JS, Englund JA, File TM, Fry AM, et al. Clinical Practice Guidelines by the Infectious Diseases Society of America: 2018 Update on Diagnosis, Treatment, Chemoprophylaxis, and Institutional Outbreak Management of Seasonal Influenza. Clin Infect Dis. 2019;68(6):895–902.

